# Contraceptive congruence: acknowledging pregnancy ambivalence in a novel measure of contraceptive use

**DOI:** 10.1101/2023.09.21.23295857

**Authors:** Hallie N. Nelson, Erik Lehman, Sarah Horvath, Cynthia H. Chuang

## Abstract

**Background:** The historical view that pregnancy intention is dichotomous (i.e., intending or not intending pregnancy), and the notion that all individuals not intending pregnancy should be using highly effective contraceptive methods, oversimplifies how we view contraceptive decision-making. To better understand this, we studied contraceptive congruence as an alternative, 3-level measure describing methods as very congruent, somewhat congruent, or incongruent with one’s individual attitudes about becoming pregnant.

**Methods:** Secondary data analysis included 982 MyNewOptions study participants who were not intending pregnancy within the next year. The cross-sectional survey assessed attitudes about how important it is to avoid pregnancy, how pleased/upset one would be if pregnant, and current contraceptive method use. Participant answers to attitudinal questions and effectiveness of current contraceptive method were used to determine congruence categories.

**Results:** Contraceptive methods included LARC (8%), other prescription methods (50%), non-prescription methods (30%), and no method (12%). Methods for 23% of participants were very congruent, 48% somewhat congruent, and 29% incongruent with attitudes about becoming pregnant. Contraceptive congruence was significantly associated with contraceptive satisfaction in bivariate analysis. Predictors of contraceptive congruence included being married or living with partner, full-time employment, and intending future pregnancy in the next 1-5 years.

**Conclusion:** Contraceptive congruence is a novel measure that acknowledges pregnancy ambivalence and is associated with higher contraceptive satisfaction scores. Future contraception research should strive for robust, patient-centered measures of contraceptive use that acknowledge the complex attitudes affecting individual contraceptive behavior and satisfaction.

---

**What is already known on this topic:** Pregnancy intention is historically seen as a dichotomous outcome (“intended” vs “unintended”), does not capture the full breadth of pregnancy ambivalence, and may be an oversimplification of an individual’s reality.

**What this study adds:** To better understand this complex interaction of pregnancy intention with contraceptive use, we propose contraceptive congruence as an alternative, 3-level measure that describes methods as very congruent, somewhat congruent, or incongruent with one’s individual attitudes about becoming pregnant. Contraceptive congruence is a novel patient centered measure that acknowledges pregnancy ambivalence, is associated with higher contraceptive satisfaction scores, and with further research could change contraceptive counseling in the future.

**How this study might affect research, practice or policy:** The differences observed in contraceptive congruence and contraceptive effectiveness suggests that better strategies are needed to measure patient-centered contraceptive method use. Clinical practice and policy must advance how we use reproductive life planning, shared decision making, motivational interviewing, and patient autonomy to help patients find contraceptive methods that are aligned with their pregnancy intentions, ambivalence, and individual preferences.

## 1. Introduction

With effectiveness of over 99%, long-acting reversible contraception (LARC), which include the intrauterine device (IUD) and the contraceptive implant, are often touted as the preferred methods for reducing unintended pregnancies.^1–3^ However, indiscriminate promotion of LARCs is problematic for several reasons, including the inaccurate assumption that pregnancy intention is dichotomous – i.e., either intending pregnancy or not intending pregnancy. On the contrary, many individuals feel some degree of ambivalence even when stating they are or are not intending pregnancy.

Commonly used measures of pregnancy intention have generally assessed whether participants want/intend to have a baby in the future.^4^ Although a “not sure” category is often offered and used by researchers to define ambivalence, we suspect that these “not sure” responses are not capturing the full breadth of pregnancy ambivalence. In contraceptive behavior studies, individuals who self-report not intending pregnancy are often expected or encouraged to use a LARC or other highly effective form of contraception. While efficacy is a convenient and easy outcome measure, it is also flawed.

Misunderstanding pregnancy ambivalence and its role in contraceptive decision making is not only an issue for research methodology, but a critically important concept that needs to be better understood to provide patient-centered clinical care. Gubrium and colleagues state that no one contraceptive method should be first line for everyone and furthermore endorsing LARC methods for everyone may be perpetuated by the Healthy People 2020 goal of reducing method failure from 12.4% to 9.9%.^5 6^ In a recent study that supports individualizing contraceptive counseling, participants desiring to avoid pregnancy had higher odds of any and more consistent contraceptive use, but not use of a more effective method.^7^ To acknowledge these complexities in contraceptive decision-making, we need to include patient-centered perspectives in how we define whether contraceptive choices are congruent with individual situations, instead of always advocating for highest efficacy methods for all.

Our objective is to propose an alternative 3-level measure, contraceptive congruence, that categorizes the contraceptive method an individual is using as being very congruent, somewhat congruent, or incongruent with their attitudes toward future pregnancy. We aim to define this new research measure and describe its association with contraceptive method satisfaction, as well as the traditional hierarchy of contraceptive method effectiveness using data from the MyNewOptions study. We hypothesize that contraceptive congruence will yield different results than method effectiveness alone.

## 2. Methods

### 2.1. Study sample

Data for these cross-sectional analyses are from the baseline survey (prior to study interventions) of the MyNewOptions randomized controlled trial. The parent study tested the effectiveness of web-based interventions to assist insured self-identified women in making individualized contraceptive choices (ClinicalTrials.gov identifier:).^8^ Study participants were members of Highmark Health plans in Pennsylvania and sampled from the Highmark enrollee database. Eligible self-identified women were aged 18-40, not intending pregnancy in the next 12 months, not surgically sterile (or current partner with vasectomy), sexually active with reported male partner(s), and had Internet access. The baseline survey, completed in 2014, assessed pregnancy and contraceptive history, current contraceptive use and behaviors, and sociodemographic factors. Further details regarding the MyNewOptions study protocol are previously published.^9^

There were 987 participants in the MyNewOptions study; the analytic sample for the current analysis was limited to the 982 participants who provided complete survey responses for the main independent and dependent variables of interest, as described below (2.2 Measures). The MyNewOptions study was approved by the Institutional Review Board.

### 2.2. Measures

#### 2.2.1. Outcome Variable: Contraceptive Congruence with Pregnancy Attitudes

Contraceptive congruence is a 3-level variable describing whether an individual’s contraceptive method is very congruent (VC), somewhat congruent (SC), or incongruent (IC) with one’s attitudes about getting pregnant. The research metric variable was constructed based on the participant’s responses to three questions—current primary contraceptive method, importance of avoiding pregnancy, and pleased/upset if became pregnant. Contraceptive methods were categorized into four groups representing highest to lowest effectiveness: 1) long-acting reversible contraceptive (LARC) including intrauterine devices (IUDs) and contraceptive implants, which have a failure rate of <1%, 2) other prescription methods (i.e., oral contraceptives, injectable, patch, vaginal ring), which have a typical failure rate of 6-9%, 3) non-prescription methods (i.e., barrier methods, spermicide, withdrawal, fertility-awareness based methods (FABM)), with a typical failure rate of 12-24%, and 4) no method. Importance of avoiding pregnancy was assessed with the question, “Thinking about your life right now, how important is it to you to avoid becoming pregnant?”—the response choices were very important, somewhat important, a little important, and not at all important. How pleased/upset one would be if pregnant was assessed with the item, “Now, imagine that you have just found out today that you are pregnant. How would you feel?”—the response choices were very upset, somewhat upset, unsure, somewhat pleased, and very pleased.

The definitions for very congruent, somewhat congruent, or incongruent are shown in Figure 1. Whether a participant’s contraception is VC, SC, or IC is dependent on individual responses to the pregnancy attitude questions. The strictest definition for congruence applies to participants for whom it is very important to avoid getting pregnant or would be very upset if pregnant, in which case only contraceptive methods with highest effectiveness (i.e., LARC) are defined as very congruent. In this same scenario, other prescription methods would be somewhat congruent, and non-prescription or no method would be incongruent. In cases where it is somewhat/a little important to avoid pregnancy or would only be a little upset/not sure/or a little pleased if pregnant, a more moderate definition for congruence applies, where any prescription method is very congruent, any non-prescription method is somewhat congruent, and no method is incongruent. Finally, for participants stating it is not at all important to avoid pregnancy or would be very pleased if pregnant within the next year, LARCs are considered somewhat congruent, given dependence on clinicians for insertion/removal, and all other methods (or no method) are considered very congruent.

**Figure 1:**
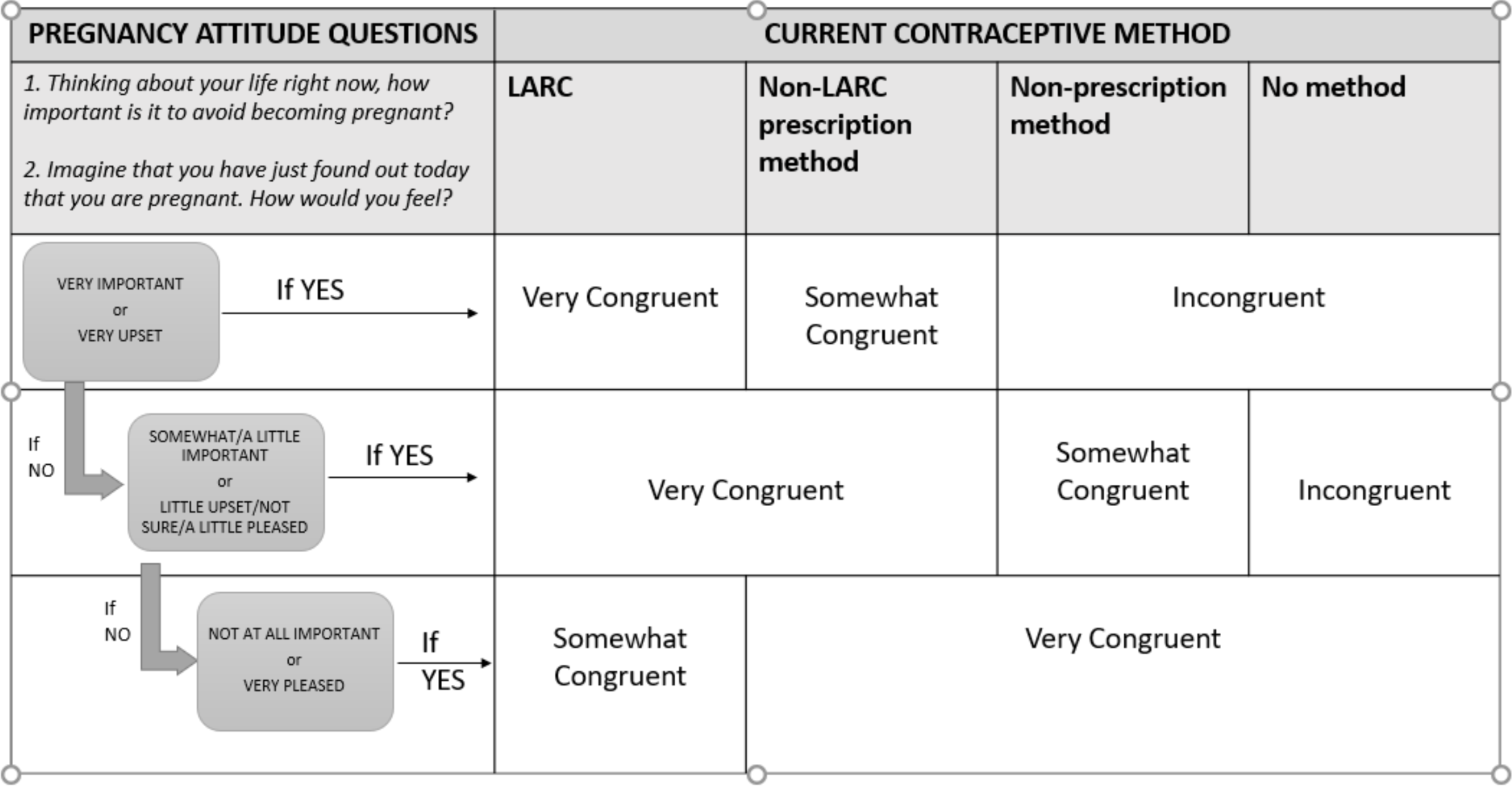
Defining Contraception Congruence.

#### 2.2.2. Independent variables

Independent variables were included that were expected to be associated with contraceptive use, including intended timing of future pregnancy, pregnancy history, and sociodemographic variables. Intended timing of future pregnancy was a 5-level variable that captured when individuals intended a future pregnancy (in 1-2 years, 2-5 years, 5 or more years, never, or not sure); only participants not intending pregnancy in the next year were eligible for the parent study. Pregnancy history variables included number of previous live births and any prior unintended pregnancy or abortion. Sociodemographic characteristics included age group, relationship status, self-reported race/ethnicity, prior education, employment status, annual household income, and religion.

The MyNewOptions baseline survey also included a single-item measure of contraceptive satisfaction: “My overall satisfaction with my birth control method(s) is…” The response options were very satisfied, somewhat satisfied, neither satisfied nor dissatisfied, somewhat dissatisfied, and strongly dissatisfied.” For analysis, contraceptive satisfaction was dichotomized as very satisfied vs. all other categories.^10^ We included contraceptive satisfaction to evaluate whether it correlated with contraceptive congruence. However, we did not include contraceptive satisfaction in the adjusted analysis, as we did not hypothesize that contraceptive congruence would be independent of contraceptive satisfaction.

#### 2.2.3. Statistical analysis

Prior to any analysis, frequencies and percentages were used to summarize all independent and outcome variables. A series of Chi-square tests were performed to assess bivariate associations between the contraceptive congruence variable and the demographic variables (Table 1). All associations with a p-value less than 0.05 were considered significant. A multivariable logistic regression was performed including the significant variables from the bivariate analysis (Table 2), except satisfaction as outlined above (2.2.2. Independent variables). We attempted to use ordinal logistic regression, but the model failed the assumption of proportional odds. Therefore, we collapsed “Somewhat Congruent” and “Incongruent” categories into one category taken as a reference group against “Very Congruent” and applied binomial logistic regression. Prior to any multivariable modeling, we checked for multicollinearity between factors using variance inflation factor (VIF) statistics. Any factors with VIF statistics >5 would be excluded from the model, but no multicollinearity was found. Odds ratios were used to quantify the magnitude and direction of any significant associations. The Hosmer and Lemeshow goodness-of-fit test was used to assess the fit of the model which was good (p=0.551). The statistical analysis software SAS version 9.4 was used to perform all analyses (SAS Institute, Cary, NC, USA).

**Table 1:**
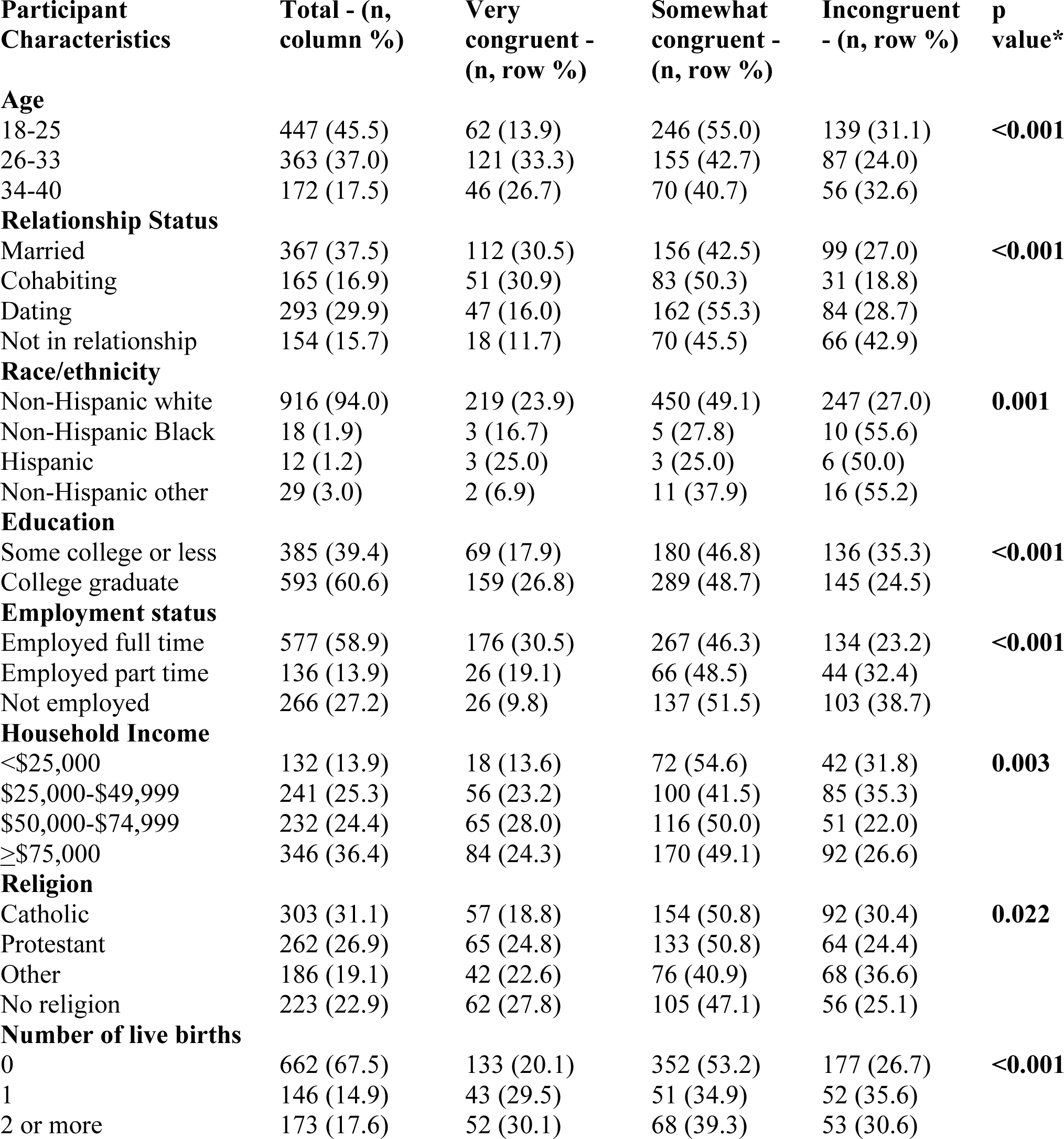
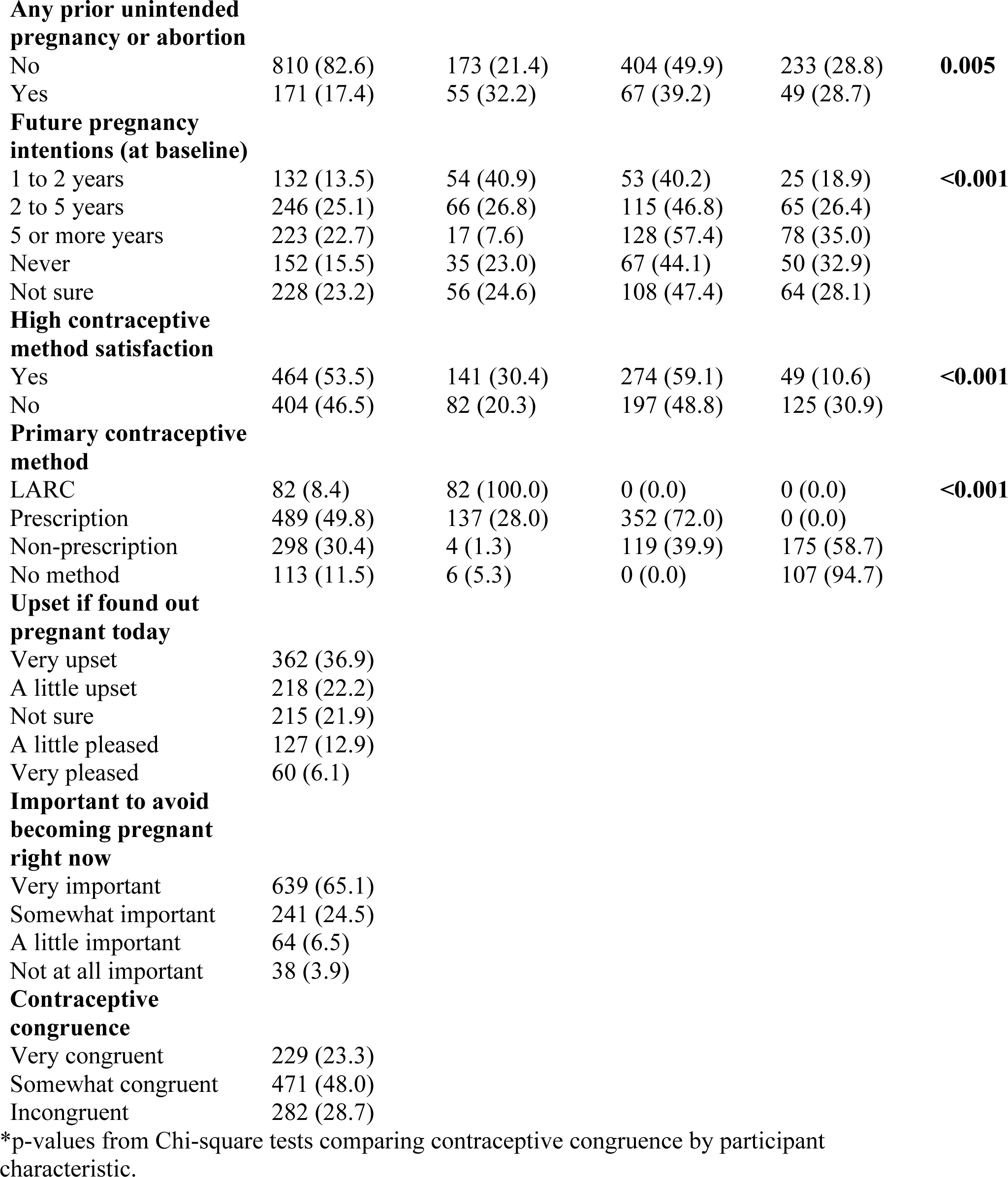
Baseline characteristics of total study sample, and by contraceptive congruence groups, n=982.

| **Participant Characteristics** | **Total - (n, column %)** | **Very congruent - (n, row %)** | **Somewhat congruent - (n, row %)** | **Incongruent - (n, row %)** | **p value\*** |
| --- | --- | --- | --- | --- | --- |
| **Age** |  |  |  |  |  |
| 18-25 | 447 (45.5) | 62 (13.9) | 246 (55.0) | 139 (31.1) | **<0.001** |
| 26-33 | 363 (37.0) | 121 (33.3) | 155 (42.7) | 87 (24.0) |  |
| 34-40 | 172 (17.5) | 46 (26.7) | 70 (40.7) | 56 (32.6) |  |
| **Relationship Status** |  |  |  |  |  |
| Married | 367 (37.5) | 112 (30.5) | 156 (42.5) | 99 (27.0) | **<0.001** |
| Cohabiting | 165 (16.9) | 51 (30.9) | 83 (50.3) | 31 (18.8) |  |
| Dating | 293 (29.9) | 47 (16.0) | 162 (55.3) | 84 (28.7) |  |
| Not in relationship | 154 (15.7) | 18 (11.7) | 70 (45.5) | 66 (42.9) |  |
| **Race/ethnicity** |  |  |  |  |  |
| Non-Hispanic white | 916 (94.0) | 219 (23.9) | 450 (49.1) | 247 (27.0) | **0.001** |
| Non-Hispanic Black | 18 (1.9) | 3 (16.7) | 5 (27.8) | 10 (55.6) |  |
| Hispanic | 12 (1.2) | 3 (25.0) | 3 (25.0) | 6 (50.0) |  |
| Non-Hispanic other | 29 (3.0) | 2 (6.9) | 11 (37.9) | 16 (55.2) |  |
| **Education** |  |  |  |  |  |
| Some college or less | 385 (39.4) | 69 (17.9) | 180 (46.8) | 136 (35.3) | **<0.001** |
| College graduate | 593 (60.6) | 159 (26.8) | 289 (48.7) | 145 (24.5) |  |
| **Employment status** |  |  |  |  |  |
| Employed full time | 577 (58.9) | 176 (30.5) | 267 (46.3) | 134 (23.2) | **<0.001** |
| Employed part time | 136 (13.9) | 26 (19.1) | 66 (48.5) | 44 (32.4) |  |
| Not employed | 266 (27.2) | 26 (9.8) | 137 (51.5) | 103 (38.7) |  |
| **Household Income** |  |  |  |  |  |
| <\$25,000 | 132 (13.9) | 18 (13.6) | 72 (54.6) | 42 (31.8) | **0.003** |
| \$25,000-\$49,999 | 241 (25.3) | 56 (23.2) | 100 (41.5) | 85 (35.3) | |
| \$50,000-\$74,999 | 232 (24.4) | 65 (28.0) | 116 (50.0) | 51 (22.0) | |
| ≥\$75,000 | 346 (36.4) | 84 (24.3) | 170 (49.1) | 92 (26.6) | |
| **Religion** |  |  |  |  |  |
| Catholic | 303 (31.1) | 57 (18.8) | 154 (50.8) | 92 (30.4) | **0.022** |
| Protestant | 262 (26.9) | 65 (24.8) | 133 (50.8) | 64 (24.4) |  |
| Other | 186 (19.1) | 42 (22.6) | 76 (40.9) | 68 (36.6) |  |
| No religion | 223 (22.9) | 62 (27.8) | 105 (47.1) | 56 (25.1) |  |
| **Number of live births** |  |  |  |  |  |
| 0 | 662 (67.5) | 133 (20.1) | 352 (53.2) | 177 (26.7) | **<0.001** |
| 1 | 146 (14.9) | 43 (29.5) | 51 (34.9) | 52 (35.6) |  |
| 2 or more | 173 (17.6) | 52 (30.1) | 68 (39.3) | 53 (30.6) |  |
| **Any prior unintended pregnancy or abortion** |  |  |  |  |  |
| No | 810 (82.6) | 173 (21.4) | 404 (49.9) | 233 (28.8) | **0.005** |
| Yes | 171 (17.4) | 55 (32.2) | 67 (39.2) | 49 (28.7) |  |
| **Future pregnancy intentions (at baseline)** |  |  |  |  |  |
| 1 to 2 years | 132 (13.5) | 54 (40.9) | 53 (40.2) | 25 (18.9) | **<0.001** |
| 2 to 5 years | 246 (25.1) | 66 (26.8) | 115 (46.8) | 65 (26.4) |  |
| 5 or more years | 223 (22.7) | 17 (7.6) | 128 (57.4) | 78 (35.0) |  |
| Never | 152 (15.5) | 35 (23.0) | 67 (44.1) | 50 (32.9) |  |
| Not sure | 228 (23.2) | 56 (24.6) | 108 (47.4) | 64 (28.1) |  |
| **High contraceptive method satisfaction** |  |  |  |  |  |
| Yes | 464 (53.5) | 141 (30.4) | 274 (59.1) | 49 (10.6) | **<0.001** |
| No | 404 (46.5) | 82 (20.3) | 197 (48.8) | 125 (30.9) |  |
| **Primary contraceptive method** |  |  |  |  |  |
| LARC | 82 (8.4) | 82 (100.0) | 0 (0.0) | 0 (0.0) | **<0.001** |
| Prescription | 489 (49.8) | 137 (28.0) | 352 (72.0) | 0 (0.0) |  |
| Non-prescription | 298 (30.4) | 4 (1.3) | 119 (39.9) | 175 (58.7) |  |
| No method | 113 (11.5) | 6 (5.3) | 0 (0.0) | 107 (94.7) |  |

**Upset if found out pregnant today**
|  |  |
| --- | --- |
| Very upset | 362 (36.9) |
| A little upset | 218 (22.2) |
| Not sure | 215 (21.9) |
| A little pleased | 127 (12.9) |
| Very pleased | 60 (6.1) |

**Important to avoid becoming pregnant right now**
|  |  |
| --- | --- |
| Very important | 639 (65.1) |
| Somewhat important | 241 (24.5) |
| A little important | 64 (6.5) |
| Not at all important | 38 (3.9) |

**Contraceptive congruence**
|  |  |
| --- | --- |
| Very congruent | 229 (23.3) |
| Somewhat congruent | 471 (48.0) |
| Incongruent | 282 (28.7) |

\*p-values from Chi-square tests comparing contraceptive congruence by participant characteristic.

**Table 2:**
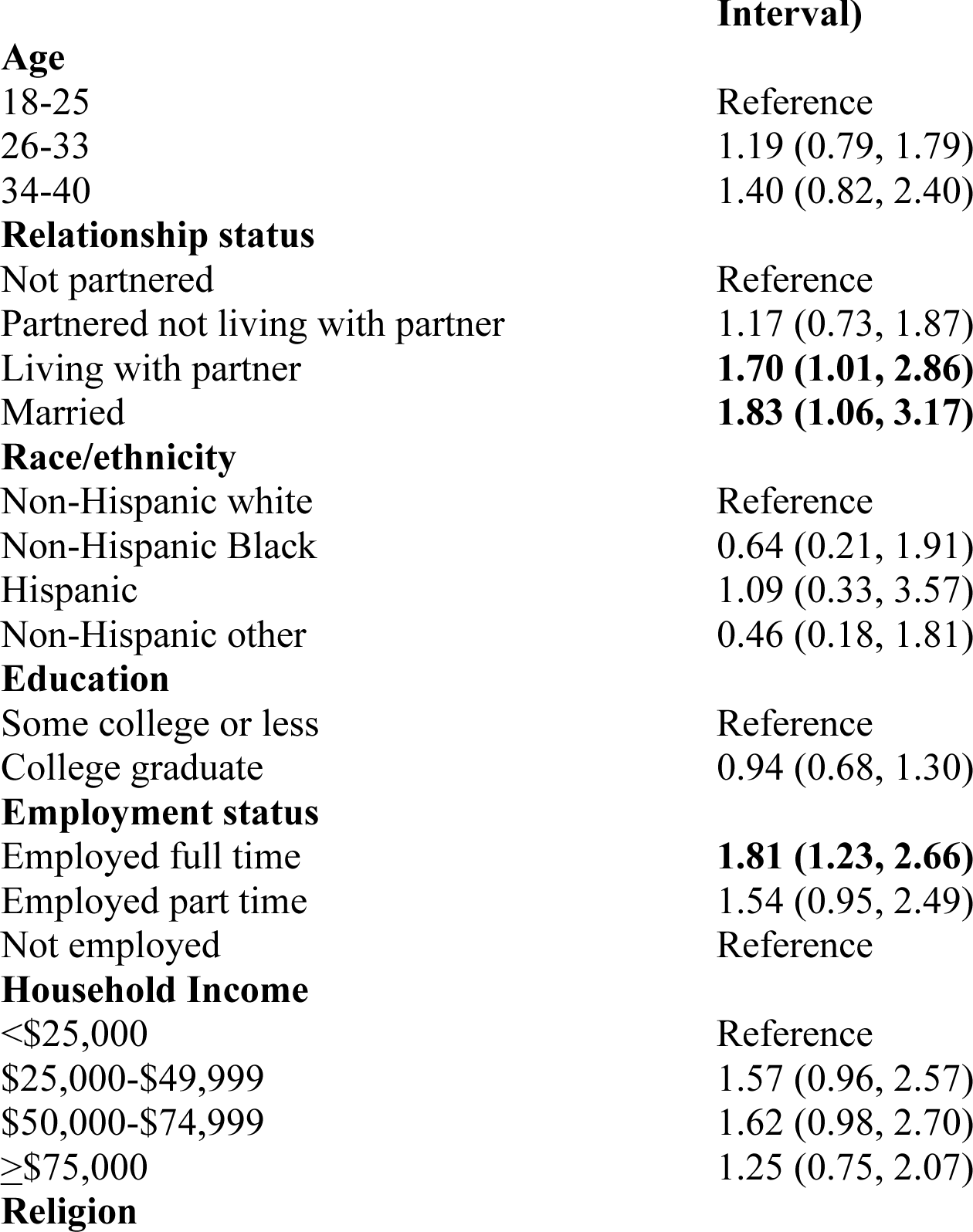
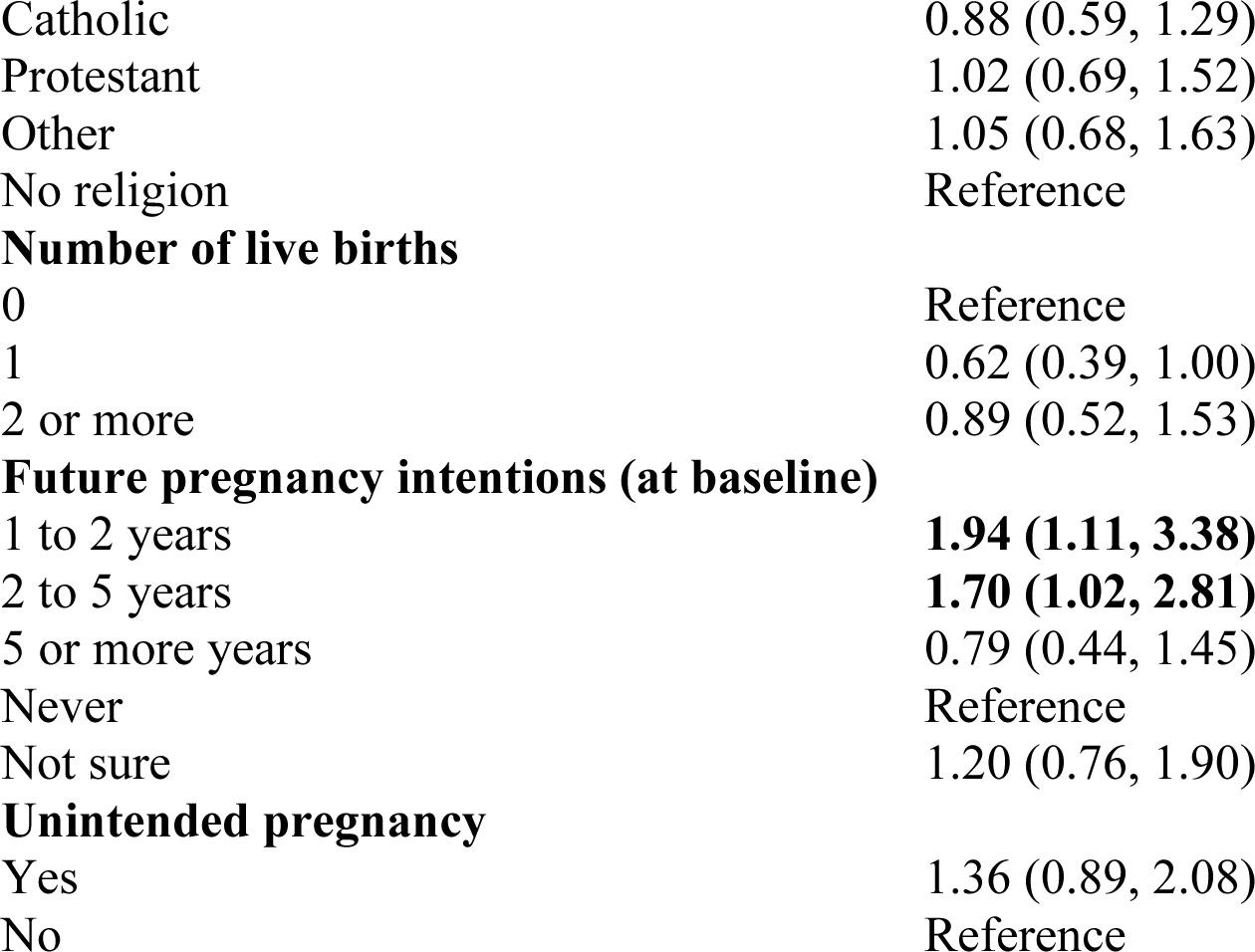
Logistic regression modeling very congruent contraceptive method use, n=934. Participant Characteristics Adjusted Odds Ratio (95% Confidence.

| **Participant Characteristics** | **Adjusted Odds Ratio (95% Confidence Interval)** |
| --- | --- |
| **Age** |  |
| 18-25 | Reference |
| 26-33 | 1.19 (0.79, 1.79) |
| 34-40 | 1.40 (0.82, 2.40) |
| **Relationship status** |  |
| Not partnered | Reference |
| Partnered not living with partner | 1.17 (0.73, 1.87) |
| Living with partner | **1.70 (1.01, 2.86)** |
| Married | **1.83 (1.06, 3.17)** |
| **Race/ethnicity** |  |
| Non-Hispanic white | Reference |
| Non-Hispanic Black | 0.64 (0.21, 1.91) |
| Hispanic | 1.09 (0.33, 3.57) |
| Non-Hispanic other | 0.46 (0.18, 1.81) |
| **Education** |  |
| Some college or less | Reference |
| College graduate | 0.94 (0.68, 1.30) |
| **Employment status** |  |
| Employed full time | **1.81 (1.23, 2.66)** |
| Employed part time | 1.54 (0.95, 2.49) |
| Not employed | Reference |
| **Household Income** |  |
| <\$25,000 | Reference |
| \$25,000-\$49,999 | 1.57 (0.96, 2.57) |
| \$50,000-\$74,999 | 1.62 (0.98, 2.70) |
| ≥\$75,000 | 1.25 (0.75, 2.07) |
| **Religion** |  |
| Catholic | 0.88 (0.59, 1.29) |
| Protestant | 1.02 (0.69, 1.52) |
| Other | 1.05 (0.68, 1.63) |
| No religion | Reference |
| **Number of live births** |  |
| 0 | Reference |
| 1 | 0.62 (0.39, 1.00) |
| 2 or more | 0.89 (0.52, 1.53) |
| **Future pregnancy intentions (at baseline)** |  |
| 1 to 2 years | **1.94 (1.11, 3.38)** |
| 2 to 5 years | **1.70 (1.02, 2.81)** |
| 5 or more years | 0.79 (0.44, 1.45) |
| Never | Reference |
| Not sure | 1.20 (0.76, 1.90) |
| **Unintended pregnancy** |  |
| Yes | 1.36 (0.89, 2.08) |
| No | Reference |

## 3. Results

The majority of the 982 participants in the analytic sample was between 18 and 25 years old, in a relationship, white, a college graduate, employed, and had an annual household income of greater than $50,000 (Table 1). Most individuals had no prior live births andwere intending to have a baby sometime in the future, with the intended timing of future pregnancy well distributed. Individuals were most likely to report they would be very upset if found out they were pregnant today (36.9%) and considered avoiding pregnancy right now to be very important (65.1%). Current contraceptive methods included LARCs (8.4%), other prescription methods (49.8%), non-prescription methods (30.4%), and no method (11.5%). The main outcome variable, contraceptive congruence, was distributed as follows: 23.3% very congruent (n=229), 48.0% somewhat congruent (n=471), and 28.7% incongruent (n=282).

Contraceptive congruence varied by participant characteristics (Table 1). Age was significantly associated with congruence, with participants aged 26-33 and 34-40 years old more likely to be very congruent (33.3% and 26.7%, respectively) compared with only 13.9% of 18-25-year-old individuals (p<0.001). Being married or cohabiting, a college graduate, employed full-time, intending pregnancy within the next 5 years, prior live birth, higher income, and prior unintended pregnancy or abortion were associated with higher contraceptive congruence.

Although the multi-level race/ethnicity and religion variables were significant in chi-square analyses (p=0.001 and p=0.022, respectively), there was not a clear trend of higher/lower congruence with any specific race/ethnicity or religious group. Congruence was associated with contraceptive satisfaction – 63% of very congruent participants had high method satisfaction, compared with 58% of somewhat congruent and 28% of incongruent participants.

Multivariable logistic regression model results are shown in Table 2, modeling very congruent contraceptive use. Compared to those who were not partnered, individuals who were living with their partner or married had greater odds of using a contraceptive method that was very congruent with their pregnancy attitudes. Likewise, full-time employment was associated with higher congruence. Finally, individuals who intend pregnancy in 1-2 or 2-5 years were more likely to be congruent than participants who never intend to get pregnant.

## 4. Discussion

In this paper, we propose a new variable, contraceptive congruence, to describe whether an individual’s primary contraceptive method is aligned with one’s attitudes toward future pregnancy to illustrate the pitfalls of using pregnancy intention alone in making assumptions about the “correctness” of one’s contraceptive method use. In this cohort of insured reproductive-age participants not intending pregnancy in the next year, only a minority (8.4%) was using a highly effective LARC, but a higher proportion (23.3%) were using a method that was very congruent with their pregnancy attitudes. While we would hope to see more individuals with high contraceptive congruence, our novel research metric suggests that measuring congruence is very different than just evaluating LARC use alone.

While we acknowledged the importance of future pregnancy ambivalence by using this novel contraceptive congruence variable, we also recognize that our methods are flawed since measurement of ambivalence has not been standardized nor validated. As described in previous qualitative studies and perspectives, pregnancy intention is not binary (“unintended” vs “intended”). Intention is much more nuanced and better described along a continuum, as suggested by many participants responding with ambivalence over pregnancy intention.^1 7 11–17^ Individuals who have strong intentions to avoid pregnancy are more likely to choose a LARC method over individuals with some ambivalence towards pregnancy, who may view a LARC as too permanent.^14^ By incorporating pregnancy ambivalence, the contraceptive congruence measure recognizes that the most effective methods are not always the most patient-centered for all individuals seeking contraception.

Measures of pregnancy intention, which can be assessed with a singular question or batteries of questions,^16 18–23^ have not been found to be durable over time. More abstract questions (e.g., lifetime intention) are less reliable, but if asked on a shorter timeframe (e.g., one year) with more specific examples, participants have a better ability to predict their intentions.^18 21^ Our data showed similar results, with individuals intending to become pregnant in the next 1-5 years being more likely to be very congruent with their method than those who never intend pregnancy. The literature also suggests that intention can be unstable when asked prospectively and retrospectively, and likely not durable over time.^24–26^ Our findings are consistent with this fluidity in intention, as participants who do not intend pregnancy ever or for at least 5 years, are half as likely to be very congruent between attitudinal questions and contraceptive method.

These results suggest that an individual’s contraceptive preferences may be better understood when their future intention is supplemented with the two additional pregnancy attitude questions, which suggests that more work needs to be done to develop a better metric. This measure can be used to describe whether contraceptive methods are aligned with individual needs, and this metric can then be used for both research measurement and clinical assessment.

Our study has several limitations. By design, our study population only included privately insured self-reported women from a single state, yielding a demographically and socioeconomically homogeneous sample. Thus, our study results are not generalizable to more racially and economically diverse populations in other parts of the U.S. However, LARC use in our study was similar to that reported in the National Survey of Family Growth, suggesting that the contraceptive use patterns may not be dissimilar from a U.S. population-based sample.^27^ Of note, this was a secondary analysis from a randomized trial testing the comparative effectiveness of web-based reproductive life planning interventions on contraceptive decision making.^9^ Because this analysis used survey data from the baseline measurement prior to randomization, the study interventions would not have interfered with the baseline participant responses. We also acknowledge the limitations of the congruence variable, which may not account for all factors that would be necessary for a robust, patient-centered measure.

Although our primary focus was to understand the implications of contraceptive use measurement for research purposes, there are implications for clinical practice and policy. Clinicians and public health agencies may encourage LARC methods to satisfy population-based goals of reducing unintended pregnancies, without acknowledging that individuals are making contraceptive choices that are influenced by many factors beyond effectiveness alone and may be driven by ambivalence over pregnancy intention and planning.^14 16 28^ Clinical practice and policy must also advance how we use reproductive life planning, shared decision making, motivational interviewing, and patient autonomy to help patients find contraceptive methods that are aligned with their individual pregnancy intentions, ambivalence, and preferences.^1 5 14 18 19 29^

By incorporating two attitudinal questions, we acknowledge pregnancy ambivalence using a measure that describes how congruent a contraceptive method is with an individual’s desire to avoid pregnancy. The differences observed in contraceptive congruence and contraceptive effectiveness suggests that better strategies are needed to measure patient-centered contraceptive method use.

## Data Availability

All data produced in the present study are available upon reasonable request to the authors

## Acknowledgement

The authors thank Highmark Health for their assistance with participant recruitment. The findings and conclusions presented are solely those of the authors and do not represent the views of Highmark Health.

